# HOW TO INTRODUCE A NEW TB VACCINE IN ADOLESCENTS AND ADULTS: Insights from Key Stakeholders in Mozambique, Southern Africa

**DOI:** 10.64898/2026.05.10.26352803

**Authors:** Agostinho Viana Lima, Doyoon Kim, Sozinho Acácio, Quinhas Fernandes, Benedita José, Benjamin Lopman, Alberto L. García-Basteiro, Kristin N. Nelson

## Abstract

Tuberculosis (TB) remains a major global health challenge, particularly in low- and middle-income countries such as Mozambique. To address this burden, promising new preventive TB vaccines targeting adolescents and adults are currently in phase III efficacy trials. This study aimed to assess stakeholders’ perspectives on priority high-risk groups, the challenges in reaching them, and potential strategies for delivering a TB vaccine. We conducted a qualitative study using semi-structured interviews with members of the National TB Program, the National Immunization Program, and the National Immunization Technical Advisory Group. Data were collected between March and July 2024. Our findings suggest that a TB vaccine program in Mozambique should prioritize individuals with comorbidities, especially those living with HIV or diabetes, and close contacts of TB patients, followed by healthcare workers, miners, and incarcerated populations. Although uptake is expected to vary across groups, relatively high coverage was anticipated among people living with HIV, TB contacts, and older adults, as well as healthcare workers, incarcerated individuals, formal miners, and in-school adolescents. To improve uptake, campaign-based strategies using mobile brigades were considered promising approaches to expand coverage. Stakeholder perspectives highlight the importance of prioritizing high-risk groups and adopting context-specific delivery strategies to support the effective introduction of a TB vaccine in Mozambique.

**Clinical trial number:** not applicable.

## BACKGROUND

Tuberculosis (TB) remains a major global health challenge, disproportionately affecting low- and middle-income countries (LMICs), particularly those in Asia and Africa. In 2025 alone, an estimated 10.7 million people developed TB, and approximately 1.23 million died from the disease (1). Evidence suggests that the burden of TB is worsened by persistent challenges in timely diagnosis and effective care. Consequently, there is an urgent need for rapid, appropriate, and effective preventive tools for TB. Addressing this need is essential to achieving the World Health Organization’s (WHO) global health goals of ending the TB epidemic by 2035, which emphasize the introduction of a new, effective vaccines is a pivotal intervention in reducing the global TB burden (2).

To date, only one vaccine - the Bacille Calmette–Guérin (BCG) - has ever been licensed for TB, but protection is variable by setting and wanes by adolescence (3–5). This is particularly evident in high-burden settings, such as Africa, where the force of infection is greater, leading to more frequent breakthrough cases (1,5). Several promising TB vaccine candidates aimed at preventing TB disease in adolescents and adults, are currently undergoing efficacy evaluation in phase III clinical trials (6). Among these, the M72/AS01E subunit vaccine candidate has shown particularly encouraging results (4,6). In a phase IIb proof-of-concept trial, M72/AS01E demonstrated approximately 50% protection against progression to active pulmonary TB over a three-year period among HIV-negative adults with latent *M. tuberculosis* infection. The vaccine is currently being evaluated in a large phase III clinical trial involving approximately 20,000 participants to further assess its efficacy in adults and older adolescents with *M. tuberculosis* infection, as well as its safety and immunogenicity in adults and adolescents without prior exposure to *M. tuberculosis* and among people living with HIV (4). Another TB vaccine candidate currently in phase III trials is MTBVAC, evaluated through the IMAGINE trial, a large-scale study assessing its safety and efficacy in preventing active pulmonary TB in children, adolescents (aged 12-18 years old) and adults (aged 18-64 years). In phase II studies, MTBVAC demonstrated a safety and reactogenicity profile comparable to BCG, with higher immunogenicity (4,7).

As these candidate TB vaccines approach pivotal efficacy trials, it is crucial to develop strategies for introducing vaccines for adolescents and adults into immunisation programmes that traditionally focus on children. This is particularly important because successful rollout will require coordination between historically vertical TB and immunization programmes and must navigate a context shaped by post–COVID-19 vaccine hesitancy, as well as the availability of existing TB preventive therapy (TPT). Currently, TB preventive therapy (TPT), is offered to individuals at high risk of developing active TB, including children under five, people living with HIV (PLHIV), and those receiving immunosuppressive therapy. Shorter rifamycin-based regimens have been shown to be as effective as six months of isoniazid, with improved safety and higher treatment completion rates (8). However, estimates from 2019 indicate that only 33% of children under five years of age worldwide who were household contacts of TB patients received preventive therapy, highlighting persistent institutional, socioeconomic, and cultural barriers. In this regard, a study reported that caregivers faced significant psychosocial challenges when incorporating TPT administration into their children’s daily routines, including financial instability, loss of social support, and stigma, factors that should be considered when planning and introducing a TB vaccine.

This study explored key stakeholders’ perspectives on introducing a potential TB vaccine, including priority high-risk groups, barriers to reaching them, and delivery strategies.

## METHODS

### Study setting and design

This study was conducted in Mozambique, a southern African country, with an estimated population of approximately 34.7 million inhabitants in 2024 (9). In the same year, the national health system comprised 1,878 health facilities, including 1,805 primary-level facilities, 58 secondary-level facilities, seven tertiary-level facilities, and eight quaternary-level facilities. However, access to health services remains a challenge, particularly in rural areas, where the average distance to a health facility is 12 km, exceeding the 10 km limit recommended by the World Health Organization (WHO) (10). Similar to other countries in southern Africa, Mozambique is among the 10 countries with a high burden of tuberculosis (TB), TB/HIV co-infection, and multidrug-resistant TB (MDR-TB). The country currently has an estimated TB incidence rate of 361 cases per 100,000 population (11) and TB is a frequent cause of death, particularly among PLHIV (12,13).

In this study, we employed an interpretative qualitative design using semi-structured interviews, which consisted of open-ended, exploratory research questions, aiming to explore key-stakeholders’ perspectives on optimal TB vaccine delivery strategies (Supplement 1). Interpretative qualitative studies seek to understand how people interpret, construct, or make meaning from their world and their experiences (14).

### Participants and sampling

For the purposes of this study, we recruited key stakeholders, divided into two subgroups: TB experts (including National TB Program staff, physicians, and researchers) and immunization experts (including Expanded Programme on Immunization staff and members of the National Immunization Technical Advisory Group [NITAG]). Participants were purposively recruited using a snowball sampling strategy and professional networks. Initially, a formal letter was sent to the Ministry of Health, specifically to the Department of Public Health, which oversees both the National TB Program and the Immunization Program, outlining the aims of the study and inviting representatives from both central and provincial levels to participate. The Ministry subsequently shared a list of potential participants and organized an online session where the research team presented the study objectives. Individuals who expressed interest and met the eligibility criteria, such as knowledge of local TB epidemiology, TB or immunization programmes, and at least two years of relevant work experience, were then registered.

In addition, researchers, physicians, and members of the NITAG were recruited based on their professional links with the research team. For these individuals, official invitation letters were also sent by email, inviting them to participate in the study.

### Data collection and procedures

Data were collected between from March to July 2024, through semi-structured interviews (Supplement 1), which were guided by open-ended questions covering specific themes designed for two groups: TB experts and immunization specialists. For example, questions related to high-risk TB groups and priority populations within these groups were directed to the first group (personnel from the National TB Program, researchers, and physicians). In contrast, questions concerning challenges in reaching vaccine target populations (particularly adolescents and adults), expected TB vaccine coverage among high-risk groups, and effective strategies for vaccine delivery were addressed to personnel from the Expanded Immunization Program and members of the NITAG.

Interviews were conducted both online (via Zoom Video Communications, Inc.) and in person, following ethical approval from CISM’s IRB and according to participants’ preferences and comfort. For online interviews, an invitation email was sent to potential participants. Upon receiving a positive response, the informed consent form was shared, signed, and returned to research team, typically one day before the interview. For in-person interviews, participants received an invitation letter together with the informed consent form. Once participation was confirmed, the date, time, and location of the interview were arranged. Before starting each interview, verbal consent was obtained to record the session. Online interviews were recorded using Zoom’s built-in recording function, while in-person interviews were recorded with a Sony ICD-PX470 digital recorder. All interviews were conducted in Portuguese by a member of the research team (AL) based in Mozambique and lasted between 30 and 80 minutes.

### Data management

#### Transcription and storage

All recordings and other study-related documents, whether physical or digital, were securely stored on the CISM server and in locked cabinets, with access restricted to the study team. The recordings were verbatim transcribed and translated from Portuguese into English using AI transcription software (Trint), and the outputs were subsequently reviewed and verified by a research assistant based in Mozambique (MM). To protect participants’ identities, each recording and transcript was assigned a unique code comprising the study acronym (TBVacMoD), site, interview type, department or area (e.g., NTP - National TB Program, IPP – Immunization Program Personnel; MHO – Ministry of Health Official, NPHD – National Public Health Specialist; ICT – TB Investigator or Clinician), and recruitment order. To further ensure confidentiality, only the last two elements - the department or area and the recruitment order - were used to identify participants in the study.

#### Data quality control

To ensure quality control, each recording and transcript was independently reviewed by two additional research team members from Emory University. Any discrepancies or inconsistencies were resolved through discussion with a third reviewer.

#### Database creation

Content data analysis was conducted using an inductive-deductive approach, enabling the identification of both emerging themes and those within the pre-defined categories. A code book was generated combing main topics, codes and sub-code (Table 1).

**Table 1.** Code Mapping.

| TOPICS | CODES | SUB-CODES |
| --- | --- | --- |
| TB Risk Groups | Demographics | Age, Sex |
|  | Socioeconomic status (SES) | Income, Education |
|  | Clinical factors | Comorbidities, Immunosuppression |
|  | Environmental factors | Household or occupational exposure (living or working conditions) |
|  | Perception of risk variation<br>(Beliefs about who is most at risk) | Perceived vulnerability |
| **TB Vaccine Priority** | Demographics | Age, Sex |
|  | Socioeconomic status (SES) | Children, Adolescents, Adults |
|  | Clinical factors | Policy decisions |
|  | Environmental factors | Perceived importance |
| **Coverage of TB Prevention** | Demographics | Clinical risk groups |
|  | Socioeconomic status (SES) | Demographic groups |
|  | Clinical factors | Socioeconomic groups |
|  | Environmental factors | Household or Work exposure (Living or Working conditions) |
|  | Compared to TPT | Perceived effectiveness and reach |
|  | Compared to SARS-CoV-2 vaccine | Perceived effectiveness and reach |
| **Challenges of Vaccination (Reaching target groups)** | Demographic barriers |  |
|  | Clinical barriers |  |
|  | Socioeconomic barriers |  |
|  | Environmental barriers |  |
| **Vaccine Delivery Strategies** | Current vaccine delivery | Routine (Fixed post) |
|  |  | Outreach brigades |
|  | Effective delivery approach | Most Effective |
|  |  | Less Effective |
|  | Dosing schedule | Number and timing of doses |
|  | Lessons from SARS-CoV-2 vaccine | Strategies used during COVID-19 |

From this analysis, two matrices were created: one quantitative, summarizing frequencies and the number of respondents, and one qualitative, containing participants’ quotes. All matrices followed the same coding structure, with participant IDs in the columns and codes in the rows.

#### Quantitative database

In the quantitative analysis, Microsoft Excel was used to record the number of participants whose responses corresponded to each predefined category of interview questions. All interview questions were reviewed by participant group, and those selected for analysis were used to identify trends relevant to the study objectives.

Each selected question was mapped to a specific research domain, including demographic characteristics, TB risk groups, and expected TB vaccine coverage. Responses to variables of interest were quantified and summarized. For demographic characteristics and TB risk groups, the frequency of key terms was extracted from interview transcripts and aggregated at the participant level. For expected TB vaccine coverage, ranges of expected coverage across risk groups were compiled from individual responses following the exclusion of outliers.

#### Qualitative database

For the qualitative analysis matrix, Microsoft Excel was used to organize the data. Relevant quotations from each participant were entered into the corresponding cells, ensuring that each excerpt was aligned with its specific code or sub-code (as illustrated in the Table 1). Each code corresponded to a particular research question, such as TB risk group categories, while sub-codes captured related attributes, including demographic characteristics, socioeconomic status, occupation, clinical conditions, and environmental factors.

Finally, a summary of key insights was created for each code and related sub-code, with illustrations using in-text citations. The presentation of findings complied with the Consolidated Criteria for Reporting Qualitative Research (COREQ) guidelines (15).

## RESULTS

### Participants’ characteristics

A total of 22 stakeholders were recruited for this study, all with expertise in TB care and immunization programs and working at different levels of the health system, including central, provincial, and district levels (Table 2).

**Table 2.** Characteristics of participants interviewed.

|  | n | (%) |
| --- | --- | --- |
| **Program affiliation** |  |  |
| TB program | 13 | (60%) |
| Immunization program | 9 | (40%) |
| **Level** |  |  |
| Central | 9 | (41%) |
| Provincial | 8 | (36%) |
| Local/District | 5 | (23%) |

### Perceptions of groups at high risk of TB in Mozambique

The respondents identified three categories of populations at high risk of TB: (i) individuals more susceptible to developing TB, (ii) individuals more likely to be exposed to and contract TB, and (iii) individuals likely to both contract and develop TB.

When asked to identify populations at high risk of TB, the majority (n=14/22) of participants initially highlighted individuals with comorbidities, particularly those living with HIV. Several participants also identified children and older adults as being at increased risk of developing TB, largely due to compromised immune function.

> “Well, in terms of high-risk groups, what I’ve noticed is that people living with HIV make up the largest number of people who develop tuberculosis within the program. In our statistics, we’ve observed that a significant number of patients are co-infected with HIV. […] children under five are particularly susceptible to developing the disease quickly, especially if they live with someone with tuberculosis in their household (ICT 04)
>
> “Regarding people with comorbidities, we are at forty-two percent (42%) TB-HIV co-infection, if I am not mistaken. That was last year. So, one of the most frequent comorbidities is indeed HIV, tuberculosis associated with HIV. We also have some cases of diabetes mellitus” (NTP, 02)
>
> “The risk groups that are already known, what are the risk groups for tuberculosis, which would be children and the elderly, immunosuppressed patients, living with HIV and other immunosuppression” (NPHS 21).

The second category emerging from participants’ accounts comprised individuals whose occupations or living conditions increase their likelihood of exposure to TB, including healthcare workers, miners, TB contacts and incarcerated populations. According to many participants, people in prison (n=13/22) face similarly overcrowded and confined conditions that heighten the risk of infection, healthcare workers (n=10/22) are routinely exposed to TB patients, and miners (n=9/22) experience prolonged exposure to silica dust and poorly ventilated environments that facilitate transmission.

> “The notification rate for health workers tends to be three or four times higher than the national notification rate. So the risk is extremely high in this population group, taking into account the number of health workers with tuberculosis versus the universe of these patients. Just to cite an example. Another group should not be left out are miners and ex-miners. With the greatest emphasis on ex-miners because of the complications resulting from exposure to silica dust in the mines” (NPHS 02).

The third category encompasses individuals from low-income households, whom many respondents (n=11/22) identified as being at high risk of both contracting and developing TB, including both drug-sensitive and multidrug-resistant forms of the disease. One participant attributed this vulnerability to limited literacy and access to information, suggesting that TB disproportionately affects economically disadvantaged and less-educated populations, particularly in rural and peripheral areas.

> “Taking into account the socioeconomic standard, […] those in the lower household, for me, are probably the most at risk of contracting the disease because they are poorly conditioned. We speak of living in houses that are poorly ventilated because we know that these diseases are transmitted with the aerosol. […] the houses […] [of] the people who have few financial conditions […] are a bit small, that don’t ventilate, […] don’t have windows there, and we have someone with the disease can easily transmit it. We have the malnutrition factor, because people who have little money and financial conditions are low, can hardly have a good meal and are unable to exercise, maintain their body, for example, pay for the gym or for something other than the gym. So, those people who have a low socioeconomic situation are in the most vulnerable situation, compared to those who are in a medium or high socioeconomic situation” (ICT 11).

In contrast, another respondent noted that multidrug-resistant TB was more prevalent among individuals in peripheral and urban settings, indicating that multidrug-resistant TB disproportionately affect populations residing in peripheral regions.

Now, with regard to the distribution of risk groups over these areas, at the moment it would be very difficult for me to look at each of them and be able to distribute them according to whether they are rural or urban, but the majority of our patients are from rural areas. Most of our patients are from rural areas in general. But when we look at tuberculosis, which is very resistant to drugs, for example, we have a higher concentration in urban and peri-urban areas” (MHO 02).

### Vaccine priority groups

When asked to identify groups that should be prioritized for a TB vaccine, two participants expressed hesitancy, suggesting that the vaccine should ideally be offered to the general population. They also warned that prioritizing specific groups could raise concerns or questions among populations that might not initially benefit from the vaccine.

> “It would be difficult, because me being a doctor, being in the clinic, I see how the child suffers. I see how this one suffers. For me personally, I would say […] it is very complicated and difficult, because it favors one against the other. Why not kids? Why not pregnant girls? Why not the old ones. Old people want to live too. But they also need a vaccine. The children are also still small, they must grow up and it’s a bit complicated to favor one group over another” (ICT 11).

Nonetheless, the majority of participants (n=12/22) expressed the view that vaccination should be prioritized for people living with HIV. They further noted that, if the vaccine were designed for this population, similar priority should ideally be extended to close contacts of TB patients, as both groups were perceived to contribute disproportionately to national TB incidence and to face particular challenges related to TB diagnosis.

> “Of all the groups I mentioned, I believe people living with HIV should be prioritized because they have a chronic condition that makes them very vulnerable. Based on epidemiological data, they have the highest morbidity and mortality rates associated with tuberculosis” (IPP 09).
>
> “Personally, I would focus on people living with HIV because they are immunocompromised and TB is harder to diagnose in this group, especially in advanced stages. So, I would prioritize people living with HIV, since they make up the largest group, around one-quarter of all TB cases” (NTP 23)

One participant encapsulated this view by proposing that TB vaccine prioritization should follow an approach similar to that adopted during the COVID-19 response, in which priority groups were defined according to their contribution to the national disease burden. This perspective, shared by many other participants, placed individuals with comorbidities, particularly those living with HIV, followed by diabetes and hypertension, as the highest priority, followed by other high-risk groups such as healthcare workers, miners, and, lastly, incarcerated populations.

> “Ok, so, basically, I would say that a strategy similar to the COVID strategy might be important. I think that with tuberculosis, one thing that is different is that we must also consider transmission. So, it is necessary to look at these groups and see which group contributes the most to tuberculosis transmission in the community. I would say it would be a good strategy to work on reducing the number of cases and significantly decreasing transmission at the community level. Clearly, I think people living with HIV would benefit from this, followed by other groups such as miners, healthcare professionals, and the incarcerated population, who would be very important” (ICT 14).

Notably, some participants (n = 6/22) emphasized the importance of prioritizing adolescents, especially those with known TB exposure, as well as older adults.

> “In addition, because tuberculosis affects more adults […] and the elderly are there, they are the majority. So, it would be important to focus on this age group” (NTP 08).
>
> “The first group to be prioritized would be children under the age of 15 […]. In the event that the caregiver has tuberculosis, close contact with the child at home is unavoidable. Home contacts would be, in my opinion, the group to take into account, for these reasons” (NPHS 02).

### Expected vaccination coverage among high-risk groups

Regarding vaccine acceptance among key high-risk groups, participants expressed differing views, suggesting that coverage would likely vary depending on perceived risk and ease of access. People living with HIV, healthcare workers, and incarcerated individuals are expected to achieve high coverage, with estimated rates of 75–95% for people living with HIV (10/22), 80– 95% for healthcare workers (9/22), and 90–95% for inmates (7/22). This is attributed both to the ease of identifying these groups within structured systems (such as healthcare facilities and prisons) and to their awareness of their elevated TB risk, which may motivate vaccine uptake.

> “[…] First of all, people living with HIV-AIDS. I expect that there will be more than 95% coverage because these people are informed. […] [also] the inmates for me it’s easier the expectation would be the same above 95% […] because […] being incarcerated or being held in prison, already reduce their ability to discuss certain interventions. They feel obliged to comply with any intervention that is brought to the prison, especially when it comes to health” (IPP 16).
>
> “Well, I think these three groups, such as (formal) miners, healthcare professionals, and the incarcerated population, are the most favored because we know where to find them. So, there are very clear and targeted strategies. I would expect coverage above ninety percent (90%)” (ICT 16).

Additionally, several population groups were perceived as likely to achieve relatively high vaccine coverage (50–80%). Two respondents perceived women as more likely to accept vaccination, as they are generally more receptive and more inclined to seek healthcare than men. However, one participant cautioned that gender norms and household power dynamics may constrain women’s ability to act on health decisions, as illustrated in the following account:

> “We had many situations where patients with tuberculosis did not reveal their condition at home. For example, a woman with tuberculosis might not inform her husband. Imagine if she does not tell her husband, how will their children take preventive tuberculosis medication if he does not know she’s sick? She might say the children did not come because they were at school, or the father took them somewhere else. However, if you delve deeper, you realize she did not reveal her illness at home; creating difficulties in ensuring the children are brought to the health facility for screening and deciding on medication. We also have situations where children live with grandparents, and the parents are far away. If the grandparent is diagnosed with tuberculosis and advised to bring the children, they might say they cannot because they are not their children” (PNT 02).

Other groups expected to reach high coverage included people with chronic conditions (such as immunocompromised individuals or those with diabetes) (5/22), TB contacts (4/22), and older adults (4/22), partly due to their frequent interactions with health services for follow-up and ongoing care.

> “Without a doubt, the vaccination coverage would be higher in women. Because women go to health units more. There’s a lot more related to women in health units, so we see more women. It’s not very common for men to go to health units. I think for women it wouldn’t be that high, I’d estimate it to be between fifty and sixty percent” (IPP 9).
>
> “[…] I would venture to say that there is more than 80% coverage. For adults, given the psychic or cognitive complexity of an adult, I can say that. So I would expect initial coverage of over 60%. […] But if we consider a group as a group from 60 years onwards, I would expect coverage of over 80%. I also mentioned the elderly over 60, because we really see that these people over 60 go to the health centers. At this age, […] these people often have other associated comorbidities […]” (IPP 16).

Moreover, formal miners (6/22) and in-school adolescents (6/22) were considered easier to reach and vaccinate, particularly through coordination with relevant institutions (e.g., ministries), compared with informal miners and out-of-school adolescents, who may be more difficult to locate. Accordingly, participants suggested that collaboration with these institutions - such as the Ministry of Energy and Natural Resources, mining companies, and other industry organizations -could be an effective strategy for reaching these groups.

> “As I said, adolescents are easy to find through coordination with the Ministry of Education. It is very easy to find them in schools, and parents easily accept it. […] our experience shows that in schools, they always accept” (IPP 18).
>
> “For artisanal miners, […] they are as if they were nomads. They don’t stop at a single point. So, it’s difficult for us to map […] this group, considering that […] if they are diagnosed up to the segment of this group of artisanal miners is difficult to locate. So, they end up being close to this segment that is difficult to map this group of miners. But, the other miners (the formal one) who are already registered, are at our levels [of reaching]” (NTP 17)
>
> “One of the strategies would be involving this association or the ministry responsible for these workers - it is the only way to reach this priority group I think because this group (informal miner) is not always present because of mobility. We might have the total number of them, but […] not all of them are here” (IPP 17).

When we quantitatively assessed estimates of vaccination coverage provided by participants, we found that respondents generally estimated that vaccination coverage would be high (80%+) among adults and adolescents, healthcare workers, and incarcerated persons, moderate (70-80%) among people living with HIV and close contacts of persons with TB, and lower (<70%) among people with non-HIV medical conditions (i.e., diabetes) and miners.

### Expected vaccination coverage relative to other TB (TPT) and vaccine interventions (COVID-19 vaccines)

For a comparison, we asked participants about the acceptability of new TB vaccines relative to recent vaccine interventions in this age group (COVID-19 vaccines) and other preventive interventions against TB (TB preventive therapy, or TPT). The majority of participants (n=14/22) believed that a TB vaccine would be highly acceptable, particularly when compared with TPT and COVID-19 vaccination. COVID-19 vaccination was associated with greater uncertainty and widespread rumors, which reduced coverage during its rollout.

> “Let me say this, regarding the COVID vaccine, when vaccination started, it came with unclear information for our population. There was a lot of fear, a lot of misinformation about the vaccine.

Unlike TB preventive therapy (TPT) - which targets only specific groups (people living with HIV and TB contacts) and requires daily or weekly medication - a TB vaccine which involved a single or two-dose regimen and be intended for a broad population. In addition, several participants noted that certain caregivers often express hesitancy about taking their children for TPT.

> I think the coverage would be greater, yes, because if we look at TPT, it is for very specific risk groups: people living with HIV and tuberculosis contacts. However, if we talk about adopting a vaccine and not focusing too much on these groups, and offering it to the general population, both those at higher risk and those at lower risk, the coverage of a new vaccine would be greater than that of TPT. So, the advantage of this would be, for example, if the vaccine were a single dose, the person gets it once, and that’s it, the vaccine will take effect. Unlike TPT, where the person has to take the medication every day, or even weekly if they are on the 3HP regimen. Therefore, I think there would be good coverage in these risk groups, considering the assumptions I just mentioned” (NTP 08).
>
> “So, the advantage of this would be, for example, if the vaccine were a single dose, the person gets it once, and that’s it, the vaccine will take effect. Unlike TPT, where the person has to take the medication every day, or even weekly if they are on the 3HP regimen. Therefore, I think there would be good coverage in these risk groups, considering the assumptions I just mentioned” (NTP 09).

On the other hands, two participants anticipated lower TB vaccine coverage, suggesting that people might prefer taking pills over receiving injections.

> “A bit, a bit complicated. I, for example, could evoke again the social aspects in which any patient prefers a pill than a vaccine. Do you understand, people are afraid of needles. If you offer the two things, people would be for a tablet that you only swallow with water than to be pricked, whether on the arm, thigh, or whatever. But whether it involves a needle or a pill, people always, the majority, would opt for the pill. So in that regard, then many people would prefer the pill TPT compared to the vaccine” (ICT 11).

### Strategies for vaccination with a new TB vaccine

Vaccination strategies were discussed primarily with immunization experts, many of whom (n=6/9) reported that campaign-based approaches combined with mobile brigades - particularly door-to-door strategies for adults and school-based delivery for adolescents - are more effective than routine services for reaching these groups, as they rarely seek care at health facilities. Nonetheless, routine vaccination should be maintained for individuals who actively seek healthcare, such as women, people with comorbidities and the elderly.

> “That might be an opportunity to give the vaccine. All of that has to be considered. Adults are very difficult to reach. Stopping work to go to a clinic for any kind of treatment, even wound care, is already hard. So, thinking of a vaccine for adults seems very complicated and unrealistic to me, unless it’s done through a campaign. You would have to go door-to-door on Saturdays. But even then, adults rarely put aside their tasks just to get a preventive vaccine. For adolescents and children, however, there are several strategies we can implement to reach those specific groups. School-based vaccination campaigns, for example, could help improve adherence. In addition, we have better outcomes when it comes to preventive regimens in these groups, unlike with adults, who might take it once and then abandon” (NTP 23).

An immunization specialist encapsulated this view by suggesting that the TB vaccine should initially be introduced through campaigns and mobile brigades, and only later integrated into routine immunization.

> “But whenever you start a new intervention and want to generate significant demand, it’s always good to begin with an intensified vaccination week, using mobile brigades, strong campaigns, and then afterward transition to routine” (IPP 18).

Furthermore, a few TB and immunization specialists (n=3/9) believed that combining campaign with a single-dose would be easier to implement than a two-dose regimen. In this regard, another participant emphasized that a campaign-based strategy could be more cost-effective if the vaccine is administered in one or two closely space doses, underscoring the importance of considering both the dosing regimen (single or multiple doses), the interval between doses, and the availability of funds for vaccine introduction efforts.

> “Now, back to your specific question, if it is a two-dose vaccine and the interval is short, for example, less than a month, people need to return for the second dose. In that case, I think it is important to start with campaigns, so we can find people and achieve good, high coverage. Now, if the vaccine is two doses but with long intervals, then organizing another campaign may be too costly” (IPP 18).

## Discussion

In this study, we analyzed stakeholders’ perspectives on strategies for introducing a potential TB vaccine. To our knowledge, stakeholder engagement on this topic has not previously been conducted in most high TB burden countries, including Mozambique, despite the fact that vaccine introduction strategies are likely to be country-specific. The few studies conducted so far (16–18) focused on broader implementation strategies and were conducted more than five years ago, during which time funding landscapes and program priorities may have changed.

Overall, participants identified immunocompromised individuals, particularly those living with HIV or diabetes, as well as children and older adults, as being at highest risk of developing TB disease. They also highlighted individuals with increased exposure due to occupation or living conditions, including healthcare workers, miners, TB contacts and incarcerated populations. Finally, people from low-income households were considered at elevated risk of both TB infection and progression to disease, including drug-sensitive and multidrug-resistant TB. While current surveillance systems collect data on the age and diabetes status of people diagnosed with TB, employment, socioeconomic, and carceral status is not routinely recorded. Therefore, engaging stakeholders to identify these high-risk groups is valuable, as this information is not routinely collected in TB surveillance systems in LMICs, including Mozambique, limiting the usefulness of existing data for planning vaccine introduction.

While a new TB vaccine would ideally be offered to all those eligible, financial, logistical, and possible supply constraints may require a prioritization strategy. Our findings suggest that individuals with comorbidities, particularly those living with HIV or diabetes, as well as close contacts of TB patients, should be prioritized for a new TB vaccine in Mozambique. This should be followed by other high-risk groups, including healthcare workers, miners, and, finally, incarcerated populations. A study in South Africa which explored possible introduction strategies for adult and adolescent TB vaccines suggested that a broad introduction in the general population of adults and adolescents would be ideal, but that if necessary, adolescents, due to their perceived high TB burden and transmission potential, followed by people living with HIV and healthcare workers, who face an elevated risk of TB disease, should be prioritized (16). Our findings are similar, highlighting the importance of prioritizing both individuals with comorbidities known to increase risk for TB and those whose occupation places them at high risk. Notably, participants in our study also listed close contacts of TB patients, particularly those under 15 years, as a key priority group in Mozambique. Although participants tended to prioritize younger children, while current TB Vaccine trials target older children. Evidence from studies in Ethiopia and Malawi, a neighbor of Mozambique, indicates that responses to TB care, such as uptake and adherence to TB preventive treatment (TPT), can vary by age, female gender, low education and income, poor patient acceptance, limited drug access, stigma, undernutrition, caregivers’ awareness, adverse drug reactions, and regimen type(19,20) For instance, children aged 2–15 years receiving the shorter 3HP TPT regimen were less likely to interrupt treatment, likely due to its brief duration. In contrast, older children on the 3HR regimen showed higher non-adherence, especially girls, those from low-income families, and urban residents. Healthcare workers also noted difficulties in encouraging asymptomatic household contacts to complete longer TPT courses. While some barriers are particularly pronounced in rural Mozambique - where long distances, high transport costs, limited options, and inadequate health services impede access (21) - these findings highlight the importance of implementing WHO recommendations to decentralize TB preventive services(20)

Furthermore, the prominence of people living with HIV as a key priority group highlights a major challenge for vaccine introduction. Although pivotal trials for M72-AS01_e_ and MTBVAC may have included people with HIV and the sample sizes were insufficient to establish efficacy in this population (22), vaccine’s efficacy in these populations, as well as among individuals with other comorbidities such as diabetes, remains unknown. Post-licensure studies to assess vaccine effectiveness in these groups will be essential to build the evidence base for their use in these groups.

Respondents reported that vaccine uptake is likely to vary by population group. People living with HIV, close contacts of people with TB, and older adults were expected to achieve relatively high coverage, partly due to their regular contact with health services for follow-up care. Likewise, healthcare workers, incarcerated individuals, formal miners, and in-school adolescents may achieve high coverage because they can be readily identified within structured settings. The South African study similarly noted that adolescents, people living with HIV, and health workers are groups that are highly accessible and likely to accept vaccination. Adolescents could be reached through existing school-based programs, similar to strategies used for human papillomavirus (HPV) vaccination, people living with HIV through their regular engagement with the health system, and via employer-facilitated vaccine programs, particularly those in high-risk occupations such as healthcare and mining (16). Although ease of access and institutional structures were emphasized in our study as facilitating acceptance among high-risk groups, vaccine programs which prioritize incarcerated populations or which involve employment-based vaccination require additional safeguards to ensure individual decision-making autonomy through transparency, clear communication, and the explicit right to refuse vaccination (23). These findings highlight the importance of collaborating with relevant institutions, such as the Ministry of Energy and Natural Resources, mining companies, and other sectoral bodies, to develop policies that promote vaccination without coercion.

In addition to institutional access and control, gender dynamics may further shape expected vaccine uptake. Our findings indicate that women may be more likely than men to accept a TB vaccine, given their greater familiarity with healthcare services. However, gender norms and household power relations may constrain their ability to act on this willingness. Evidence from a gender analysis on vaccine uptake similarly shows that inequalities, often reinforced by male religious leaders and household decision-making structures, strongly influence vaccination decisions. In contexts such as Mozambique, women frequently lack autonomy over their own health or that of their families due to systemic and sociocultural constraints (24,25). Even when women can make decisions, local norms often require male approval (25). Supporting this, USAID’s Gender Analysis for Vaccine Response toolkit reports that only 5% of married or in-union women make their own health decisions, while 78% indicate that their husband primarily decides, and 10% require permission to seek care (26). Although that analysis also identified safety concerns among some women, contrasting with our finding of generally high female acceptance, it underscores that willingness alone may not translate into uptake without addressing underlying gender power dynamics (26).

Our findings suggest that starting with campaign-based strategies combined with mobile brigades, such as door-to-door delivery for adults and school-based delivery for adolescents using a one or a closely spaced two-dose schedule, followed by integration into routine delivery, may be a cost-effective approach, as it can reach large populations within a short period. Supporting this approach, a narrative review of behavioral and social drivers and strategies to optimize vaccine acceptability in Kenya and South Africa recommended that, prior to the introduction of a TB vaccine, government and public health organizations should provide timely digital campaigns for the vaccine using diverse platforms (Facebook, Instagram, X, Reddit, YouTube, TikTok) and various media formats (film, text, etc.) (5). Such campaigns could help to address concerns related to safety, efficacy, or side effects, distrust in government or health authorities, misinformation or lack of information about the vaccine, low perceived risk of disease, religious or cultural incompatibility with vaccines, and social media use for vaccine information (5). This may be particularly important given that mass vaccination campaigns targeting adults and older adults can be challenging, as limited prior experience and potential perceptions of stigmatization may result in lower coverage (17). Furthermore, two similar studies reported that single-dose vaccines were consistently preferred by stakeholders due to ease of delivery, reduced logistical burden, and better adherence, particularly when reflecting on the poor uptake of second doses during COVID-19 (16,17). These results underscore the need to consider the dosing regimen, interval between doses, past experience with adults, sociocultural beliefs and, lastly, available funding when introducing the vaccine.

Future studies should consider perspectives beyond TB and immunization program officials such as budget and financing stakeholders, to ensure broader representation of all relevant decision-makers at the national level.

## Conclusion

In this study, we explored stakeholders’ perspectives on strategies for introducing a potential TB vaccine. While several high-risk groups were identified, our findings suggest that vaccination should focus on high risk groups: people living with HIV and close contacts of people with TB, followed by healthcare workers, miners, and incarcerated populations. Although high coverage is expected, it will likely vary depending on access and ease of accessibility of each population. In this context, campaign-based strategies using mobile brigades, door-to-door for adults and school-based for adolescents, may help improve coverage. A single-dose vaccine would likely achieve higher uptake than a two-dose regimen, particularly if followed by integration into routine delivery. However, successful implementation will depend not only on the vaccine’s clinical profile but also on how it is communicated, positioned, and integrated into social and health systems, as well as the costs involved.

## Data Availability

All data relevant to the present study are in the article. The data used for this article are of qualitative nature in which participants shared personal data/information, which is difficult to anonymize completely. Further, participants have not provided informed consent for their data to be stored in a public repository. Given these reasons and to guarantee confidentiality, we would rather not share our data in a public repository. Data are available from the authors upon reasonable request and with approval from the scientific executive committee of the Manhica Health Research Centre. Request can be submitted via

## List of abbreviations

BCG: Bacille Calmette–Guérin
CISM: Centro de Investigação em Saúde da Manhiça
COREQ: Consolidated Criteria for Reporting Qualitative Research
HIV: Human immunodeficiency virus
LMIC: Lowand Middle-Income Countries
MDR-TB: multidrug-resistant TB
NITAG: National Immunization Technical Advisory Groups
PHIV: People with HIV
TB: Tuberculosis
TPT: preventive TB treatment
SSI: Semi-structured Interviews
WHO: World Health Organization

## Declarations

### Ethics approval and consent to participate

This study was conducted according to the protocol, the Declaration of Helsinki, as well as other locally relevant regulations. Ethical approval for the study was obtained from the Manhiça

Health Research Centre’s Institutional Review Board in Mozambique (Ref. CIBS-CISM/038/2023), Written and verbal consent was obtained from all participants. Participants were informed that they could withdraw from the study or stop the interview at any time and without any implication. The interviews were audio recorded with the permission, and according to the comfort of the respondents.

### Consent for publication

No applicable.

### Availability of data and materials

All data relevant to the present study are in the article. The data used for this article are of qualitative nature in which participants shared personal data/information, which is difficult to anonymize completely. Further, participants have not provided informed consent for their data to be stored in a public repository. Given these reasons and to guarantee confidentiality, we would rather not share our data in a public repository. Data are available from the authors upon reasonable request and with approval from the scientific executive committee of the Manhiça Health Research Centre. Request can be submitted via

### Competing interests

The authors declare no competing interests.

### Funding

This publication was produced by the National Institutes of Health/National Institute for Allergy and Infectious Disease (grant number 1K01AI166093-01A1). The funder had no role in the design or conduct of the study or reporting of results.

## Authors’ contributions

**Conceptualization:** Agostinho Viana Lima, Kristin N. Nelson.

**Data curation:** Agostinho Viana Lima, Doyoon Kim, Kristin N. Nelson.

**Formal analysis** Agostinho Viana Lima, Doyoon Kim, Sozinho Acácio, Quinhas Fernandes, Benedita José, Alberto L. García-Basteiro, Kristin N. Nelson.

**Funding acquisition:** Kristin N. Nelson.

**Investigation:** Agostinho Viana Lima, Doyoon Kim, Kristin N. Nelson.

**Methodology:** Agostinho Viana Lima, Kristin N. Nelson.

**Project administration:** Agostinho Viana Lima, Kristin N. Nelson.

**Resources:** Agostinho Viana Lima, Kristin N. Nelson.

**Supervision:** Sozinho Acácio, Alberto Garcia-Basteiro, Kristin N. Nelson.

**Validation:** Agostinho Viana Lima, Doyoon Kim, Sozinho Acácio, Quinhas Fernandes, Benedita José, Alberto L. García-Basteiro, Kristin N. Nelson.

**Visualization:** Agostinho Viana Lima, Doyoon Kim, Sozinho Acácio, Quinhas Fernandes, Benedita José, Alberto L. García-Basteiro, Kristin N. Nelson.

**Writing – original draft:** Agostinho Viana Lima.

**Writing – review & editing:** Agostinho Viana Lima, Doyoon Kim, Sozinho Acácio, Quinhas Fernandes, Benedita José, Alberto L. García-Basteiro, Kristin N. Nelson.

## Acknowledgments

We are grateful for the time and effort contributed by the participants. We thank Neusa Torres and the social sciences team, field staff at the Manhiça Health Research Center, and Casey Randleman and Shamika Chavda for project and analysis support at Emory University.

## Notes

### Competing Interest Statement

The authors have declared no competing interest.

### Author Declarations

This study was conducted according to the protocol, the Declaration of Helsinki, as well as other locally relevant regulations. Ethical approval for the study was obtained from the Manhiça Health Research Centre's Institutional Review Board in Mozambique (Ref. CIBSCISM/ 038/2023)

